# Lesion Normalization and Supervised Learning in Post-Traumatic Seizure Classification with Diffusion MRI

**DOI:** 10.1101/2021.08.06.21261733

**Authors:** Md Navid Akbar, Sebastian Ruf, Marianna La Rocca, Rachael Garner, Giuseppe Barisano, Ruskin Cua, Paul Vespa, Deniz Erdoğmuş, Dominique Duncan

## Abstract

Traumatic brain injury (TBI) is a serious condition, potentially causing seizures and other lifelong disabilities. Patients who experience at least one seizure one week after TBI (late seizure) are at high risk for lifelong complications of TBI, such as post-traumatic epilepsy (PTE). Identifying which TBI patients are at risk of developing seizures remains a challenge. Although magnetic resonance imaging (MRI) methods that probe structural and functional alterations after TBI are promising for biomarker detection, physical deformations following moderate-severe TBI present problems for standard processing of neuroimaging data, complicating the search for biomarkers. In this work, we consider a prediction task to identify which TBI patients will develop late seizures, using fractional anisotropy (FA) features from white matter tracts in diffusion-weighted MRI (dMRI). To understand how best to account for brain lesions and deformations, four preprocessing strategies are applied to dMRI, including the novel application of a lesion normalization technique to dMRI. The pipeline involving the lesion normalization technique provides the best prediction performance, with a mean accuracy of 0.819 and a mean area under the curve of 0.785. Finally, following statistical analyses of selected features, we recommend the dMRI alterations of a certain white matter tract as a potential biomarker.

## 1 Introduction

Traumatic brain injury (TBI), a condition in which physical injury to the brain causes temporary or permanent impairment to brain function [18], can have far-reaching consequences [12]. A particular concern is the development of seizures in response to the injury, which can indicate a diagnosis of post-traumatic epilepsy (PTE) [9]. This work studies the development of seizures with subjects from the Epilepsy Bioinformatics Study for Antiepileptogenic Therapy (EpiBioS4Rx) which aims to identify accurate biomarkers of epileptogenesis and subsequently design and perform preclinical trials of antiepileptogenic therapies. Reliable biomarkers could then serve as surrogate endpoints for future clinical trials of antiepileptogenic therapies in humans.

Magnetic resonance imaging (MRI) methods that probe structural and functional alterations after TBI are promising for biomarker detection [14]. Diffusion-weighted MRI (dMRI) is one method that researchers have used to identify changes in white matter (WM) connectivity that relate to seizure development [10]. Many dMRI studies are based on estimating quantitative indices such as fractional anisotropy (FA), mean diffusivity (MD), and apparent diffusion coefficient (ADC), which characterize water diffusion in WM tracts [11, 20]. Of these indices, only FA is found to be a promising biomarker for epilepsy and TBI, whereas MD and ADC are not [11, 20]. As such, we focus our exploration in this paper on FA features.

FA has been used to distinguish and classify subtypes of temporal lobe epilepsy (TLE) in a region of interest (ROI) based analysis with perfect accuracy, using logistic regression [17]. Other ROI based studies have modeled both functional and structural connectivity derived from functional MRI (fMRI) and dMRI, respectively, to predict seizure outcomes in postsurgery TLE patients with 100% accuracy, using principal component analysis (PCA), correlation, and Euclidean distance [16]. FA features extracted from voxel-wise analysis have also been applied for the prediction of consciousness recovery with an 86% accuracy, using the machine learning (ML) technique of linear discriminant analysis (LDA) [20]. Even though a perfect classification is reported in both [16, 17], the subjects did not have any lesions and the imaging data in each was acquired in a single site, with the same scanner. FA features obtained from tract based spatial statistic (TBSS) have also been used to train support vector machine (SVM) ML classifiers, to characterize seizure development with 85.7% accuracy [3].

To the best of our knowledge, none of these works compensate for brain lesions and deformations that often occur with moderate-severe TBI, and that are not well corrected by the standard nonlinear registration [20].

In this work, we explore methods to address TBI induced deformation through dMRI preprocessing, including the application of lesion normalization techniques to dMRI scans obtained from multiple sites, for patients with complete follow-up data. Here we perform cost function masking [4], a technique normally used for fMRI analysis, when normalizing the dMRI into Montreal Neurological Institute (MNI) space. Once the dMRIs are in MNI space, we systematically extract FA features of different WM tracts from four parallel preprocessing pipelines, two of which utilize the proposed lesion normalization technique. We then test ML models encompassing different feature selection methods and binary classifiers to comprehensively evaluate the predictive performance of each pipeline, and statistically test the discriminative power of the features selected by the best ML model of each pipeline. Finally, we recommend the combination of the preprocessing pipeline and the ML model that yields the highest mean accuracy and mean area under curve (AUC) as a promising tool for the early prediction of seizure occurrence in TBI patients, while recommending any feature that obtains statistical significance as a potential biomarker.

## 2 Methods

### 2.1 dMRI Acquisition and Lesion Segmentation

According to the EpiBioS4Rx protocol [2], moderate-severe TBI patients with evidence of contusion were eligible for enrollment. A total of 22 patients (18 male, 4 female; average age = 45.0, SD=20.9) were chosen for this work: 13 (63%) experienced at least one seizure more than 7 days after injury, whereas the remaining 9 were seizure-free. Mean Glascow Coma Scale at emergency department arrival was 9.4 (SD=4.1) Multimodal imaging was acquired, on average, 15 days postinjury (SD=9.1). The acquired imaging included, but not limited to, different MRI sequences such as T1 MRI, dMRI, T2, and T2-weighted fluid attenuated inversion recovery (FLAIR).

This work was approved by the UCLA Institutional Review Board (IRB# 16-001 576) and the local review boards at each EpiBioS4Rx Study Group institution. Written informed consent to participate in this study was provided by the participants’ legal guardian/next of kin.

To account for brain deformations and lesions due to TBI, 3D T2-FLAIR images for each patient were manually segmented using ITK-SNAP [27]. Parenchymal contusions and brain edema were segmented together as one 3D mask for this analysis. Manual segmentations were reviewed by clinicians with expertise in neuroradiology.

### 2.2 Preprocessing Pipelines

In order to prepare the dMRI for analysis, the acquired dMRI scans are processed in the FMRIB Software Library (FSL) [23], to estimate diffusion tensor imaging (DTI) parameters and FA images. Following the preliminary registration, the subsequent steps are carried out to extract TBSS features in four different pipelines, as outlined in Fig. 1. In Pipeline 1 and 2, no lesion information is used and are labeled as non-lesion analyses (NL). Both NL pipelines use nonlinear registration, as seen in Fig. 1, to transform the FA images of individual subjects from the native space into the standard MNI space, by registering the subject image to the standard FA template (Std). In Pipeline 1, skeletonization for identifying FA values along the different WM tracts, is carried out using the mask and distance map from the Std following the method reported in [25]. In Pipeline 2, study-specific (SS) mean FA derived from the patients is used for masking individually registered FA images and skeletonization [13]. Pipelines 3 and 4 use lesion information by compensating for the T2-FLAIR lesion mask via cost function masking while registering subject space FA masks to MNI space, shown as lesion normalization (Les) in Fig. 1. Pipelines 3 and 4 extend the approach as outlined in [1]. These two Les pipelines then follow the same steps involved in the first two NL pipelines. Following skeletonization, mean FA values are calculated along a total of 63 WM tracts and WM bundles obtained from the JHU-DTI atlas [25].

**Fig. 1:**
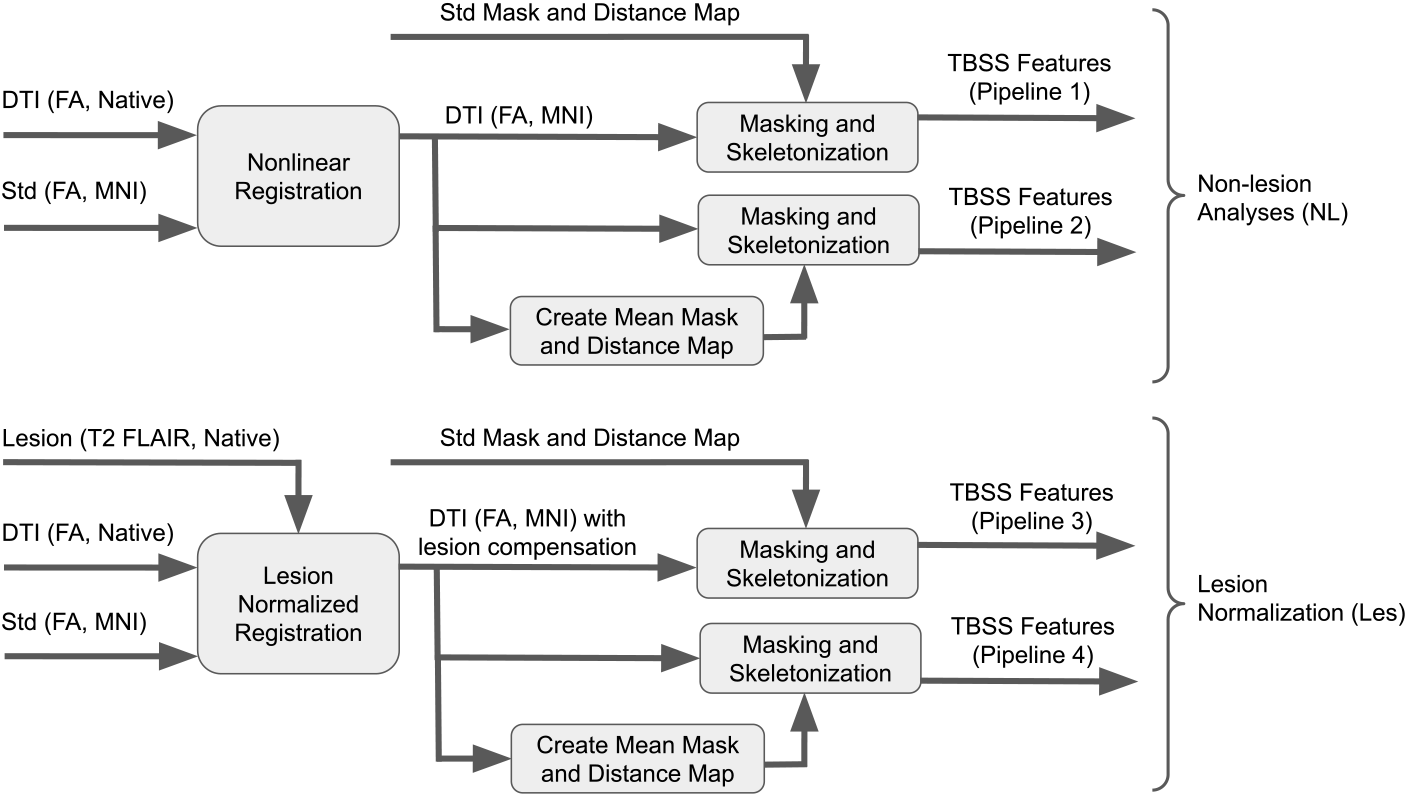
The four different TBSS preprocessing pipelines.

### 2.3 Lesion Normalization

The lesion normalization technique is based on the normalization procedure given in [1], where a native space T1-weighted scan of a pathological brain is transformed into a lesion compensated image in the MNI space using a binary mask of the lesion (1=lesion area, 0=non-lesion area), and an MNI152 structural scan template. In this work, we have modified that technique by replacing: the T1-weighted subject scan with an FA image obtained from DTI, and the MNI152 structural scan by a standard FA template in the MNI space.

To produce the lesion mask, we first transformed the lesion mask obtained from a T2-weighted fluid attenuated inversion recovery (FLAIR) image to the MNI space and then converted it to the necessary binary mask format.

### 2.4 Feature Selection and Classifiers

In classification and prediction tasks, choosing a suitable classifier depends greatly on the type of input data and the desired outcome. In this work, the mean FA, taking values between 0 and 1, along the different WM pathways form a continuous-valued numerical input matrix **X** ∈ ℝ^*N×J*^, where *N* = 22 is the number of subjects analyzed and *J* = 63 is the total number of features. The number of seizure labels (a late seizure or not) forms a binary categorical output vector **y** ∈ ℝ^*N×*1^. For this input-output pair, we conducted an exhaustive search among several binary classifiers of potential interest, namely: Adaboost, Random Forest, Gradient Boosting Machines, XGBoost, LDA, SVM, multi-layer perceptron (MLP) neural network, logistic regression, and Gaussian Naive Bayes.

The input feature dimension *J*>*N*, even without taking into consideration the reduction of training samples due to cross-validation (CV). Many of these features may add noise and hurt the overall classifier performance by increasing complexity and over-fitting [8]. It is thus recommended to select the features that will maximize the discriminative power of the classifier. Feature selection mainly takes two broad forms: wrapper methods and filter methods [8]. Wrapper methods attempt to find the optimal selection of features specifically for a given classifier, where the result of the selection is non-transferable. Conversely, filter methods dissociate the feature selection from the classifier training, by aiming to maximize some objective of similarity between the individual features and the target labels. In this work, we investigated three univariate filter methods versus no feature selection applied, on the training set of each CV fold. The univariate methods independently rank the usefulness of each feature in explaining the target label, and they are, namely: Mutual Information (MI), *χ*^2^-test, and F-test. MI between the *j*-th feature and **y** is estimated using the nearest neighbor method [21]. The *χ*^2^-test and F-test are carried out in their standard formulations as implemented in the scikit-learn library [19] of Python, where the test results are looked up in their respective tables to calculate the relevant scores.

### 2.5 Evaluation Metrics and Experimental Details

The data is divided into a 5:1 train and test set split, following a six-fold CV. Since the data is unbalanced, a randomized stratification is included in the CV strategy, such that the percentage of samples for each class is preserved in each fold. For hyperparameter tuning, a validation set is temporarily created from only the first training set. The classifiers are then trained on the resulting reduced train set. Once the parameters for the classifiers are selected, based on the preliminary performance of the classifiers on this validation set, the CV experiments are then subsequently carried out using the entire train set.

To assess the performance of each feature selection-classifier model trained on the training set in each fold, we evaluate them over the respective test set in each of those CV folds. For each model involving a feature selection technique, the number of features selected is varied from one and up to ten features, resulting in ten repetitions of the six-fold CV. The specific features chosen, in each of those ten repetitions, are essentially incorporated inside the CV procedure. Among these eleven combinations of features tested (no feature selection, and one through ten features selected), we record the number of features that lead to the best performance and the corresponding performance metrics: mean prediction accuracy, mean AUC of the receiver operating characteristic (ROC) curve, mean sensitivity, and mean specificity. The accuracy is given by

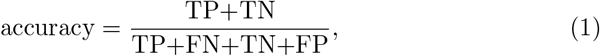

where TP, FN, TN, and FP are the total true positive, false negative, true negative, and false positive, respectively. The mean accuracy is recorded with its standard error of mean (SEM), calculated for a 95% level of confidence. The ROC analysis makes use of the 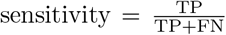, and the 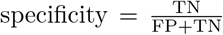. The variation of the mean ROC curve is estimated by a binomial distribution approximation [24], for a 95% level of confidence.

The best performing model (in terms of AUC, and accuracy to tie-break if necessary) for each pipeline is identified. Since the two classes are slightly imbalanced, AUC is preferred over accuracy. In order to test for potential biomarkers, the features selected in the best performing model from each pipeline are compared between the seizure and no seizure group, with the Mann Whitney U tests performed on the entire dataset. If the null hypothesis is rejected, the feature may be recommended as a potential biomarker of late seizures.

The entire analysis is run on a workstation equipped with a 9th generation core-i7 3.6 GHz CPU, 64 GB of RAM, and an RTX 2080 Ti GPU hardware. The software used are FSL 6.0.3, and Python 3.7.4 with scikit-learn version 0.23.2 and xgboost version 1.1.1.

## 3 Results and Discussion

In preprocessing, the processed FA images were registered to the standard Human Connectome Project (HCP1065) DTI FA template [26]. The HCP1065 FA is slightly bigger in volume than FSL’s FMRIB58 FA [23], and has a higher resolution than the ENIGMA DTI FA [25]. In preliminary experiments with dMRI scans of TBI patients, the HCP1065 FA yielded better classification performance compared to the other two templates, and was thus chosen as the Std.

The effect of the lesion normalized registration on the MNI space dMRI, when compared to a standard non-linear registration, can be seen in Fig. 2. Lesion normalization was observed to cause discontinuity of the WM tracts in the spatial location of the lesions, and the resulting images also appeared less deformed.

**Fig. 2:**
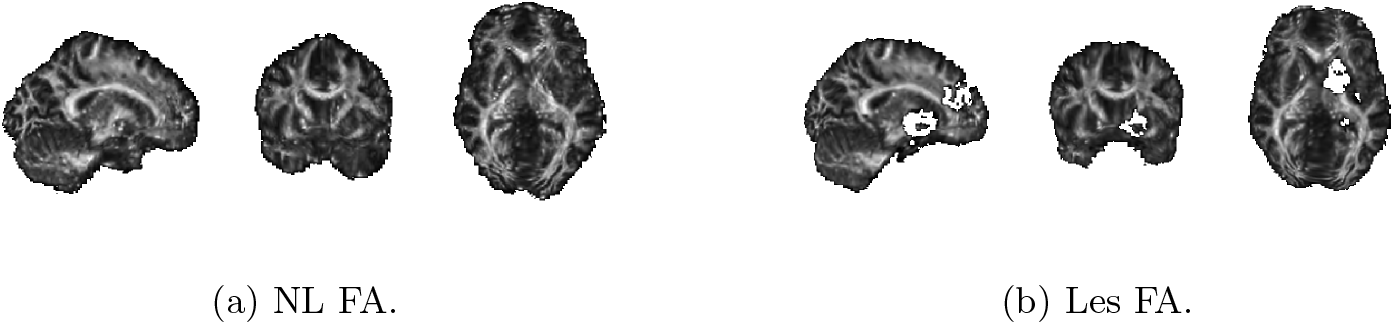
Patient 35 registered in MNI space, without and with lesion compensation.

In our six-fold CV experiments, the Adaboost classifier consistently outperformed Random Forest, Gradient Boosting Machines and XGBoost classifiers. Among the other classifiers tested, LDA beat SVM (linear kernel), tuned MLP (two hidden layers), logistic regression, and Gaussian Naive Bayes classifiers. Since Adaboost and LDA performed the best in aggregate among all classifiers tested, we report only the performances of Adaboost and LDA. The hyperparameters tuned were the number of trees and estimators in these classifiers, while all other parameters were kept at their scikit-learn and xgboost defaults. The hyperparameters were varied among values = {4,8,16,32,50,64,100}, before being tuned to 100, a value that maximized the accuracy and AUC on average in the validation set.

The Adaboost and LDA classifiers were then used in combination with all the features and the three feature selection methods, for each pipeline. Results of these experiments are recorded in Table 1. We observe that both variants of the lesion normalization outperform the respective variants of the non-lesion analysis, in terms of the best seizure prediction performance by each (shown in bold). Interestingly, both standard template registration pipelines also beat their corresponding study-specific registration pipelines in performance. In particular, the Les Std (pipeline 3) with a *χ*^2^-test based feature selection and LDA as the classifier performs the best overall. LDA emerged as the better classifier, with the best performance from each pipeline, and a faster average run-time (0.27s) compared to Adaboost (0.97s). The ROC plots for the best performing model from each pipeline (displayed in bold in Table 1) are shown in Fig. 3.

**Table 1:**
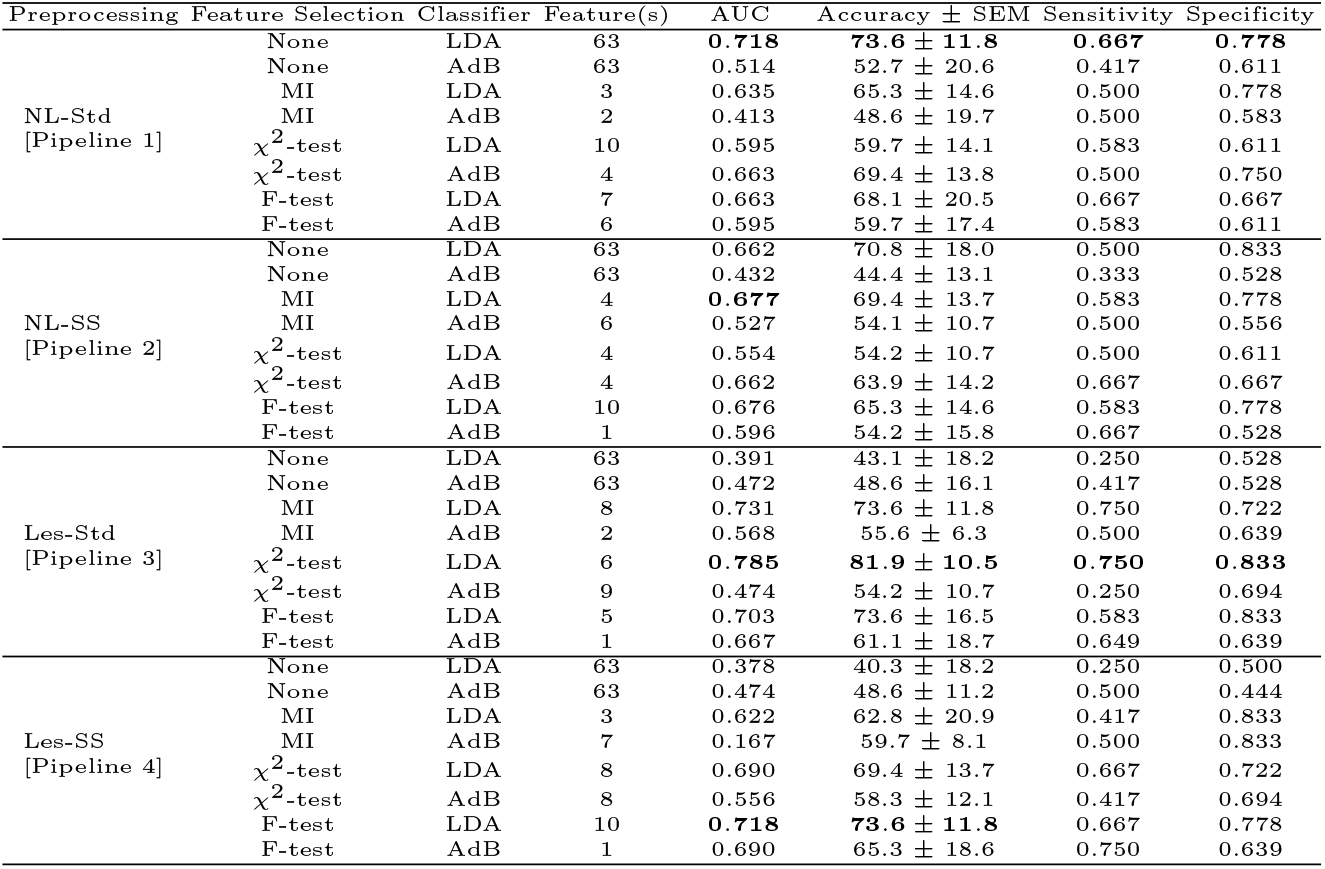
Performance comparison of the different pipelines using selected univariate feature selection methods and chosen binary classifiers.

**Fig. 3:**
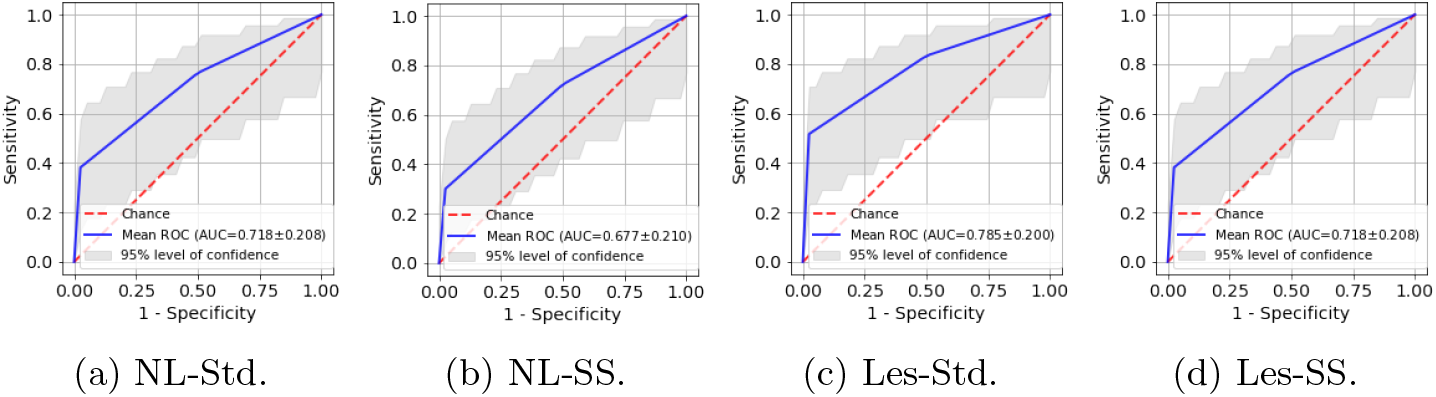
Receiver operating characteristics for the best performing models.

In order to understand whether the features that resulted in the best classification performance could also be used as potential biomarkers, we compared the distribution of the features from the best classifier in each pipeline across groups using the Mann Whitney U test. Aside from the best model in pipeline 1, each of the other models chose *k* features (where the value of *k* depended on the specific model), in each of the six folds of the CV. From the aggregate of all the six folds, we ranked the features chosen in decreasing order of their frequency of appearance. The top *k* most frequent features were then selected for comparison between the seizure and no seizure group. The features with uncorrected p-values less than 0.05 are reported in Table 2, along with their Bonferroni corrected p-values [7]. Table 2 shows that the mean FA in the anterior limb of the internal capsule right (ALIC-R) is significantly different between groups. White matter abnormalities of this tract have been reported earlier in patients with temporal lobe epilepsy [15]. These results provide further evidence that dMRI alterations in ALIC-R may be a potential biomarker of epileptogenesis after TBI. It is worth noting that alterations in other tracts (UNC-R, CST-L, IC-R, and PLIC-R) that we found to be significant before the Bonferroni correction have been shown to be associated with seizure propagation, duration and seizure-related WM abnormalities [5, 6, 22]. It may be that as the considered sample size increases, the effects of alterations in these tracts will become more evident.

**Table 2:** Comparison of features between the two seizure groups using the Mann Whitney U test, with p-values corrected using the Bonferroni correction.

| Preprocessing and Model | Features | Potential Feature* | Test Statistic | Uncorrected p-value | Corrected p-value |
| --- | --- | --- | --- | --- | --- |
| NL-Std [None,LDA,63] | 63 | FX/ST-L | 33.0 | 0.0475 | 1.000 |
|  |  | UNC-R | 30.0 | 0.0308 | 1.000 |
| NL-SS [MI,LDA,4] | 4 | FX/ST-L | 33.0 | 0.0475 | 0.1900 |
| Les-Std [$\chi^2$-test,LDA,6] | 6 | ALIC-R | **22.0** | **0.0081** | **0.0486** |
|  |  | UNC-R | 30.0 | 0.0308 | 0.1848 |
| Les-SS [F-test,LDA,10] | 10 | ALIC-R | 22.0 | 0.0081 | 0.0810 |
|  |  | CST-L | 27.0 | 0.0192 | 0.1920 |
|  |  | IC-R | 33.0 | 0.0475 | 0.4750 |
|  |  | PLIC-R | 30.0 | 0.0308 | 0.3080 |
|  |  | UNC-R | 31.0 | 0.0357 | 0.3570 |

\* FX/ST-L=Fornix/stria terminalis left, UNC-R=uncinate fasciculus right, ALIC-R=anterior limb of internal capsule right, CST-L=corticospinal tract left, IC-R=internal capsule right, PLIC-R=posterior limb of internal capsule right.

## 4 Conclusion

In this work, we introduced cost function mapping, a lesion normalization technique, to the study of dMRI and compared various preprocessing pipelines using classification performance to judge among them. From the presented experiments, we found that the pipeline involving lesion normalization and a standard template skeletonization provides the best late seizure prediction performance. Lesion normalization shows an improvement of eight percentage points in mean accuracy and seven percentage points in mean AUC, when compared to the standard template skeletonization pipeline with non-lesion analysis. Following statistical analyses of selected features, we find evidence that the dMRI alterations of ALIC-R may serve as a biomarker of seizures post TBI.

This work has certain limitations. Since this longitudinal study is ongoing, the experiments are only carried out on a limited number of patients from the cohort, for whom the complete follow-up data has been obtained. Additionally, the mean FA values used for the analyses might not adequately characterize the spatial distribution of FA values along the tracts, where additional distribution descriptors might provide more discriminative information. Furthermore, deep convolutional models, which could provide better classification performance, have not been tested due to the limited number of samples.

For future work, we aim to expand the study by including more subjects and examine the effects of lesion normalization on dMRI registration and classification in greater detail. We would also like to explore other techniques involving the analysis of white matter tracts, and investigate other neural network classifiers that may be trained to perform well with few data samples.

## Data Availability

This is an ongoing study. It will be announced later, whether the data will be made available to the public.

